# FAIR Data Cube, a FAIR data infrastructure for integrated multi-omics data analysis

**DOI:** 10.1101/2023.04.23.23289000

**Authors:** Xiaofeng Liao, Thomas H.A. Ederveen, Anna Niehues, Casper de Visser, Junda Huang, Firdaws Badmus, Cenna Doornbos, Yuliia Orlova, Purva Kulkarni, K. Joeri van der Velde, Morris A. Swertz, Martin Brandt, Alain J. van Gool, Peter A.C. ’t Hoen

## Abstract

**Motivation:** We are witnessing an enormous growth in the amount of molecular profiling (-omics) data. The integration of multi-omics data is challenging. Moreover, human multi-omics data may be privacy-sensitive and can be misused to de-anonymize and (re-)identify individuals. Hence, most biomedical data is kept in secure and protected silos. Therefore, it remains a challenge to reuse these data without infringing the privacy of the individuals from which the data were derived. Federated analysis of Findable, Accessible, Interoperable, and Reusable (FAIR) data is a privacy-preserving solution to make optimal use of these multi-omics data and transform them into actionable knowledge.

**Results:** The Netherlands X-omics Initiative is a National Roadmap Large-Scale Research Infrastructure aiming for efficient integration of data generated within X-omics and external datasets. To facilitate this, we developed the FAIR Data Cube (FDCube), which adopts and applies the FAIR principles and helps researchers to create FAIR data and metadata, to facilitate re-use of their data, and to make their data analysis workflows transparent, and in the meantime ensure data security and privacy.

## 1 Introduction

It is now widely acknowledged that understanding the mechanisms underlying health and disease requires the concerted study of different molecular levels (Deoxyribonucleic Acid (DNA), Ribonucleic Acid (RNA), proteins, metabolites). Moreover, a transition is needed from static and somewhat simplified views, to dynamic and more comprehensive views on biological pathways. Description of these pathways is usually accomplished by measuring the comprehensive assembly of molecular features in a biological system on the level of genes, transcripts, proteins and metabolites, *i.e*., the study of -omics: genomics, transcriptomics, proteomics and metabolomics. Currently, this is not simple nor scalable. As such, there is an increasing need to combine -omics data from different sources (multi-omics) in order to achieve a better understanding of biological systems, but the data and their associated metadata are not always FAIR: Findable, Accessible, Interoperable, and Reusable[1]. For that reason, the Netherlands X-omics Initiative has developed a multi-omics data infrastructure that facilitates FAIR-compliant multi-omics data storage and analysis. The proposed data infrastructure provides an analysis environment for (federated) data handling and analysis, in the meantime ensure data security and privacy.

This paper introduces our solution of integrated analysis on FAIR multi-omics data in decentralized databases. In the remainder of this paper, section 2 investigates existing work in this research direction. Section 3 presents the design and implementation of the FDCube and section 4 showcases the use of FDCube in the Trusted World of Corona (TWOC) project[2]. Finally, section 5 discusses further developments.

## 2 Related work

There are several tools that aid researchers in managing research metadata in a FAIR manner, for instance the FAIR Data Station[3], the FAIR-in-a-box[4] approach, and the DataFAIRifier[5]. Most of these tools focus on the production of FAIR data, including ingestion, generation, and publication.

For a more comprehensive coverage of FAIR processes including data management, data security, data exchange, and federated analysis, additional tools are required. For example, MOLGENIS is an open-source web-application covering the typical flow of human genomics data including data collection, management, analysis, visualization, and sharing, as well as offering support to make data FAIR[6, 7]. MOLGENIS can be hosted on-site and stores the data locally in a PostgreSQL database. This offers all the advantages of a database system including a local access control system (in light of the European General Data Protection Regulation) together with detailed data management.

The Personal Health Train (PHT)[8] concept is underlying a number of approaches for decentralised analysis of health-related data. The essence of the PHT approach is the analogy of a station representing the data source and a train representing the research question (or a computational request) visiting the data stations. Stations range from very large databases to small personal lockers containing the data of one person. Each station has its own set of house rules describing what a visiting ‘train’ is allowed to do with its data[8]. By moving trains towards stations rather than moving data, copying of data is avoided, hence data remains under complete control of the person or institute generating the data, thereby reducing privacy concerns around data sharing.

DataSHIELD[9] implements the idea of bringing algorithms to the data to ensure data privacy and security. DataSHIELD facilitates (co-)analysis of (harmonised) biomedical, healthcare and social-science data stored at one or multiple locations. The analysis requests are sent from a central analysis machine to several data-holding machines, which store the harmonised data to be co-analysed. The datasets are then analysed simultaneously, but in parallel. MOLGENIS developed a DataSHIELD implementation called Armadillo in its MOLGENIS suite.

Vantage6[10, 11] is a different implementation of the PHT concept. Vantage6 enables collaboration between multiple parties by allowing to participate in one or multiple studies across multiple data stations.

In terms of programming language, DataSHIELD restricts itself to a single language (R)[12] and to a pre-defined library of functions and algorithms. In contrast, Vantage6 allows the researcher to send a request to use their preferred programming language, as long as the language is supported by the targeted data station.

To advance and further build upon the currently available federated, FAIR solutions for the scientific community, we here present the FDCube for public use under an open MIT license. In contrast to the more generic MOLGENIS Armadillo approach, the FDCube contains more specialised services for the analysis of multi-omics data. For example, we adopt the Investigation, Study, Assay (ISA) metadata schema to capture metadata about (-omics) experiments in a hierarchical manner, in which the different omics layers can be integrated within one project and connected through common identifiers. To our best knowledge, this is the first federated infrastructure designed for multi-omics data analysis. The FDCube is developed based on the principle that data should be “as open as possible and as closed as necessary” [13]. By incorporating a FAIR Data Point (FDP) component[14], the metadata can be as open as possible and made FAIR-at-the-source. By integrating a Vantage6 component[10], the data security and privacy can be ensured during federated analysis.

In comparison to other FAIR initiatives such as CEDAR[15], FAIRDOM[16] and Omics Discovery Index (Omics Discovery Index (OmicsDI)) [17], the FDCube has a number of additional strengths. First of all, CEDAR and FAIRDOM both focus mostly on general metadata management (*i.e*., FAIRification of datasets), whereas the FDCube provides additional solutions for -omics (meta)data. In addition to metadata generation and publication, FDCube goes a step further by dealing with federated analysis tools and approaches in order to promote reusability of data. Furthermore, OmicsDI facilitates the access and dissemination of -omics datasets by indexing metadata coming from the public datasets from various resources, but it expects data in a common XML format. Because there is no use of standard ontologies it is difficult to adhere to the FAIR principles, whereas the FDC supports the use of ontologies by utlizing FAIR Data Station which combined a set of ontologies to support the metadata model based on ISA.

## 3 Result

The FDCube is a technological framework for the storage, analysis and integration of multi-omics data. The FDcube reuses and extends existing open software components/modules and initiatives. This includes the FDP[14] and Vantage6[10]. Further elements of the FDCube are the ISA metadata framework[18, 19] for capturing general study metadata, sample (including basic sample characteristics), and assay metadata, and the Phenopackets[20] standards for capturing phenotypic description of a patient/sample. The concept of the FDCube is illustrated in Fig 1 and detailed below from the perspective of a dataset owner and a researcher respectively. The complete and detailed documentation on the FDCube can also be found at https://github.com/Xomics/FAIRDataCube/wiki.

**Fig 1.**
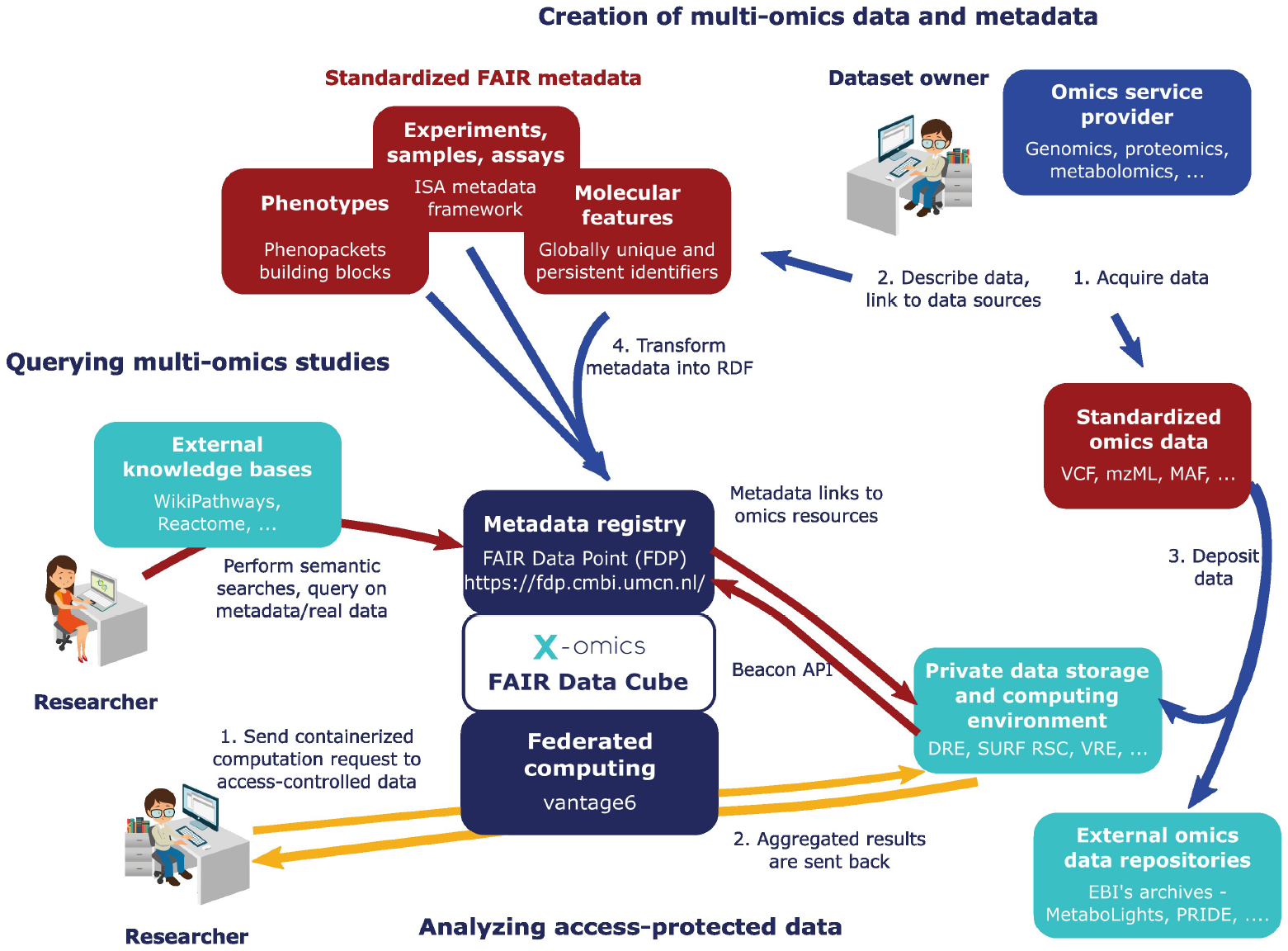
The concept of the FDCube from a dataset owner and dataset user (researcher) perspective. The dataset owner (***right upper corner***), for instance an -omics service provider, can (*1*) acquire the data and (*2*) describe the data in a standardized FAIR metadata schema. The standarized -omics data can then be (*3*) deposited in any appropriate resource/database with the links to that data included in the metadata schema. Next, the metadata schema can be (*4*) transformed into RDF to be added to a metadata registry, such as a FDP. The standardized -omics data formats can be obtained from external -omics data repositories, like MetaboLights (metabolomics), PRIDE (proteomics), and others. On the other hand, researchers (***left lower corner***) can perform semantic searches on the (publicly) shared metadata registries by querying the (multi-)omics studies published in them, either or not with the help of external knowledge bases. Alternatively, they can analyze access-protected data by (*1*) sending a containerized computation request to the data, which will then be send to the private data storage and computing environment through Vantage6. This will then (*2*) send the aggregated results back to the researcher. These aggregated results prevent (re)identification of individual samples.

### 3.1 Dataset owner

A dataset owner (Fig 1; right upper corner) acquires the dataset (*1*) and registers it (*2, 3*) by publishing the metadata on a metadata registry(*4*), such as in a FDP. The FDP is a metadata repository that provides public access to metadata in accordance with the FAIR principles[14]. The FDP helps dataset owners to publish the metadata of their dataset, and facilitates other researchers to find and access information (metadata) about these registered datasets, including pointers to the actual location of the data (which can in theory be anywhere). This is irrespective of data access restrictions and licenses, which is typically arranged by the dataset owner at the place where the data is stored.

Considering the various metadata formats adopted by the different research communities who focus on multi-omics data, it is desirable to adopt a standard metadata format as a template for submitting of study metadata. To this purpose, we employed the ISA metadata framework[18, 19] as our basic framework, to capture and standardize study (design) information from different -omics metadata schemes. The ISA metadata schema is widely adopted by a number of research communities, for example for submission of metabolomics data as implemented by EMBL’s European Bioinformatics Institute (EMBL-EBI) in their MetaboLights repository[21].

In biomedical studies, clinical characteristics and phenotypic information of study subjects may need to be collected in addition to (-omics or other) measurements data. This information is essential for making biologically-relevant interpretations from research experimental data. Thus, phenotype data need to be standardized as well, so that researchers and clinicians can more easily link these phenotypic characteristics also to other types of biomedical data. To achieve this, the Phenopackets framework[20] as developed by the Global Alliance for Genomics and Health, was adopted in the FDCube. This framework comprises a comprehensive data structure (data model), and makes use of common ontology terms, in order to categorise and connect different types of phenotype data.

### 3.2 Researcher

The researcher (Fig 1; left lower corner) can be both a data set owner and a data set consumer. As a dataset consumer, the user can search any FDP for any dataset of interest. For example, one could query a FDP part of a FDCube containing multiomics molecular study data, provided its metadata is properly ontologized. Since all metadata is represented in a linked data format, the researcher can conduct semantic searches on datasets and their corresponding study information by using the SPARQL Protocol and RDF Query Language (SPARQL) query interface. The information that can be queried is the ontologized description of, for instance: study samples and their (biological) source; sample preparation details; methods and techniques applied; (-omics) measurement and (data) analysis strategies, workflows and reports, including the detected (molecular) data features, research group affiliations, *etc*. Example questions that may be asked are:

1. Find all studies which use mass spectrometry-based metabolomics and study a specific metabolic disorder;
2. Find datasets with more than two -omics types and more than 100 individuals;
3. Find measurements for proteins and metabolites that belong to a particular metabolic pathway.

To analyze access-protected data and explore more complex research questions, the researcher can (*1*) send a computational request to a private data storage and computing environment. This is achieved by the Vantage6 component of the FDCube. If the request is accepted by the dataset owner, the (*2*) aggregated results of the computational request are calculated at the data storage side and sent back to the researcher through Vantage6. These aggregated results prevent (re)identification of individual samples.

## 4 Demonstration of FDCube in TWOC

We adopted the Trusted World of Corona (TWOC) project to demonstrate how to utilize the FDCube for integrated multi-omics federated analysis. The TWOC project aims to contribute to a more sustainable, innovative high-quality and person-oriented healthcare system. To this end, they created a platform in which humans and machines can meet based on FAIR data, protocols and algorithms.

In Fig 2, we provide an example of the creation and application of the FDCube based on a public dataset on Coronavirus disease 2019 (COVID-19) featuring multiomics patient data by Su *et al*., 2020[22], which was FAIRified as part of the TWOC project. To demonstrate the added value of data FAIRification, we integrated the multi-omics data with data on molecular pathways from another FAIR resource: WikiPathways[23], as described in detail in Fig 5. Below is an overview of the workflows for creating, filling, and using the FDCube.

**Fig 2.**
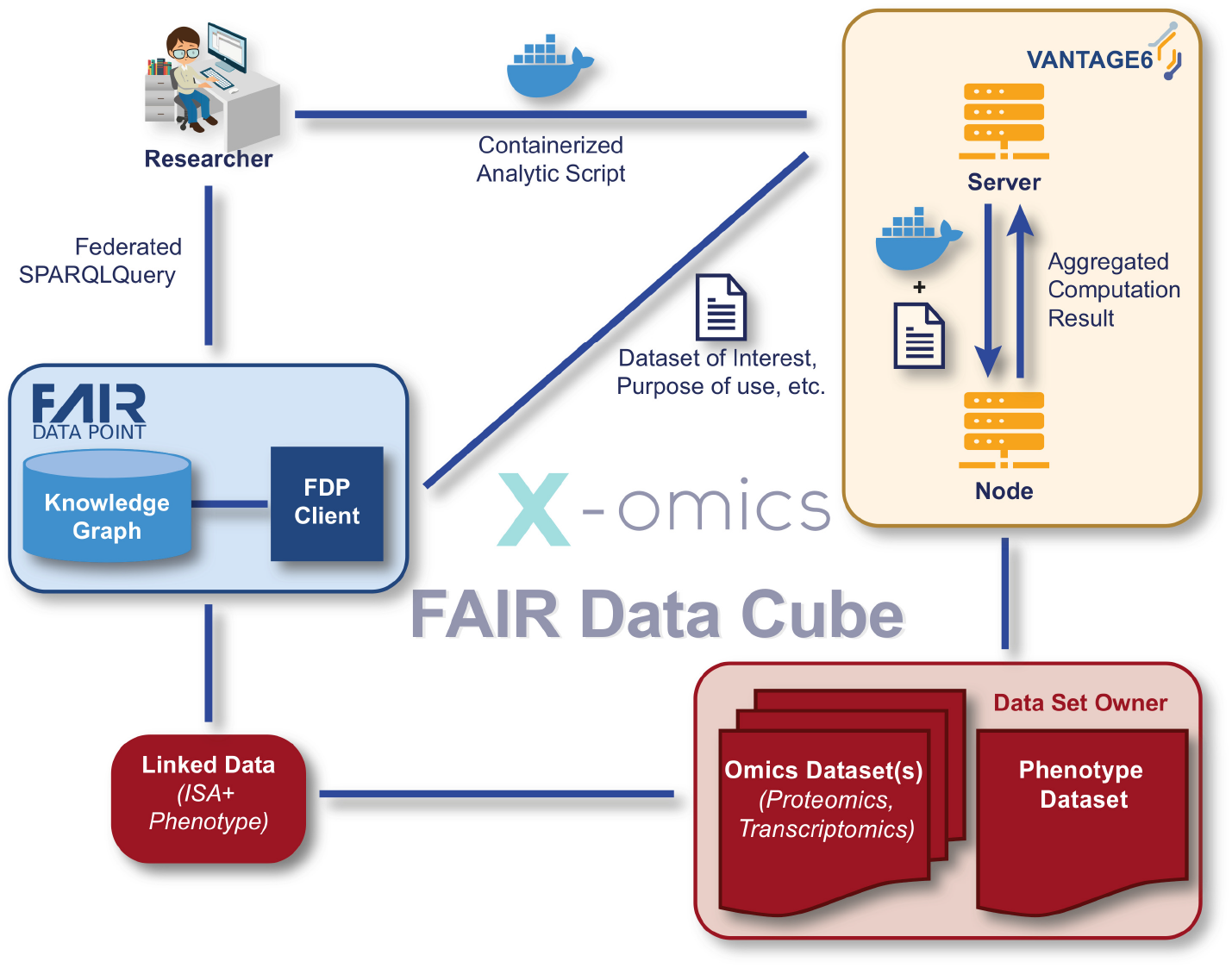
Example of how the FDCube was used in the TWOC demonstrator study. The study (meta)data, in this case publicly available data, was FAIRified through a FAIRification process into a linked data format (Turtle), and it included pointers to the actual location of the data elsewhere. This linked metadata file was stored in a FDP and could be queried through SPARQL (*blue*). The researcher (*left upper corner*) queried the data set through Vantage6 (*yellow*) for federated analysis, while the data remained at the side of the dataset owner (*red*).

### 4.1 Storage of raw and processed -omics data

A publicly available multi-modal dataset from COVID-19 patients[22] was prepared, harmonized and FAIRified as part of the TWOC project. The dataset consists of paired -omics data layers describing transcriptomics, proteomics, and metabolomics of blood samples, and includes comprehensive phenotype information (Fig 2, *in red*).

The FAIRified dataset, including documentation of the relevant (meta)data and their FAIRification processes, is publicly accessible at the TWOC’s demonstrator GitHub repository[24].

To allow interactive and joint querying of data and metadata through Vantage6 (Fig 2, *in yellow*), we store the processed -omics data along with their feature annotation files. These are both stored in a flat-text tabular .*csv* format, with features as rows and samples as columns.

### 4.2 Creation of metadata

In the TWOC project, both the ISA metadata schema and Phenopackets schema are adopted. The ISA metadata schema is used as a standard metadata schema to capture metadata about (-omics) experiments, and serializes them in an hierarchical ISA-json file using ISA tools[19, 25]. The ISA tools also provides additional functionalities to convert ISA objects into linked data file formats, for example into Turtle: a Terse RDF Triple Language file[26].

Example scripts, templates and documentation thereof are provided in our GitHub repository, in order to assist researchers in capturing study and experimental (meta)data[27]. Notably, for phenotype data, a Python script was developed based on the Phenopackets data schema, to automatically convert non-FAIRified phenotypic information into .*csv* format[27]. Furthermore, a YARRRML[28] template was written that embedded the Resource Description Framework (RDF) schema [29] of Phenopackets, by making use of the transformation service from FAIR-in-a-box[4]. This converts the .*csv* file into a linked data format. In the end, the final output with linked data, and including study and experimental (meta)data as well as phenotypic information, are uploaded into the triplestore within the FDP (Fig 2, *in blue*). This FAIRified linked data can subsequently be queried by the user through SPARQL, to extract the requested study (meta)data information.

To best assist researchers in FAIRification of their experimental (meta)data that is used as input for the FDCube, a containerized environment was created for use of the ISA-API[30], with connection to the ISA cookbook [31].

### 4.3 Querying of metadata

The FDP portal can display complete/partial metadata in a human-readable format for browsing, searching and querying of metadata. The FAIRified metadata of the TWOC demonstrator dataset was published on a FDP portal [32], as shown in Fig 3. A SPARQL query can be run against the metadata via the SPARQL query portal, to extract any requested study (meta)data information, as illustrated in Fig 4. After finding an interesting dataset via browsing or by SPARQL queries, the researcher can further run follow-up analyses on a target dataset, for example by ordering a computation request to the Vantage6 server, and if successful to retrieve the computation results from the data station via Vantage6.

**Fig 3.**
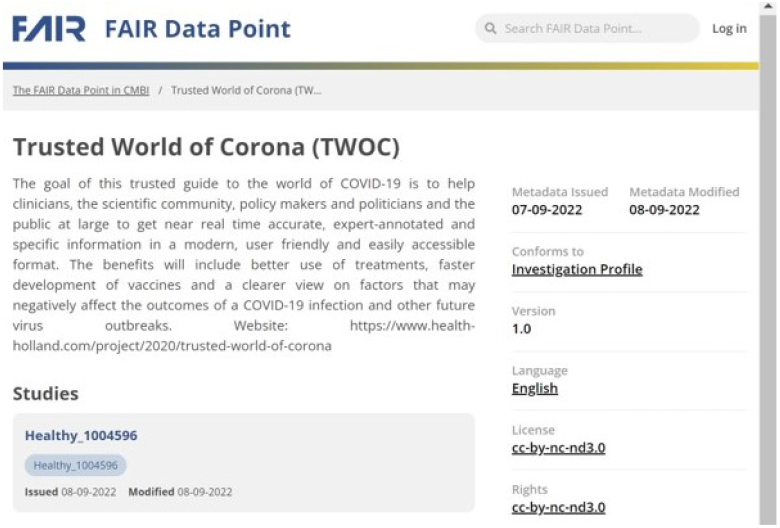
FDCube example of FAIRified metadata from the TWOC demonstrator dataset in a FDP. The figure shows a snapshot of an example study catalogue and its metadata, as published in the FAIR Data Point portal. This FAIRified metadata was generated by tooling and resources as offered within the FAIR Data Cube environment.

**Fig 4.**
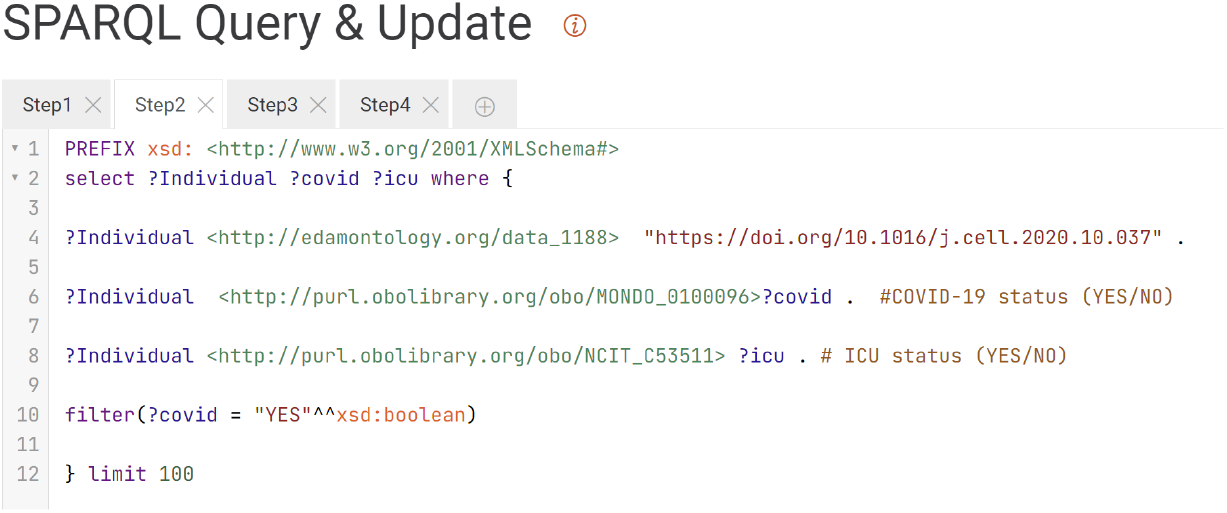
FDCube example of a SPARQL query portal in a FDP. The figure presents a snapshot of the SPARQL query portal, featuring an example query as provided by the triple store within the FDP. Both the portal and triple store are components of the FDCube environment. The displayed query corresponds to step 2 of the multi-omics data analysis described in Section 4.4. The purpose of this step is to retrieve information on individuals included in the study and to assess their COVID-19 disease status and ICU admission status.

### 4.4 Multi-omics data analysis

Together with the previous steps as described in section 4, the FDCube’s capability to support multi-omics data analysis is demonstrated in this subsection, based on the TWOC demonstrator example project. The FDCube makes use of several FAIR resources and uses pathway information collected from WikiPathways[23] to analyse transcriptomics and proteomics data from COVID-19 patients. In this example, the dataset is processed as described in 4.1. Data and code are publicly accessible at the TWOC’s GitHub repository[24, 33].

The examples consists of the following steps, as illustrated in Fig 5.

- Querying FDPs to identify relevant COVID-19 resources and their storage location.
- Fetching data of individuals participating in the study, including phenotypic information such as COVID-19 and ICU admission status.
- Obtaining subject identifiers, and using them to fetch study samples including their measurements data, as collected from the subjects.
- Retrieving experimental study group information (*i.e*., subject with COVID-19 disease, healthy control subject, ICU-admitted, and non-ICU-admitted patients) from the sample metadata in the FDP.
- Identifying a COVID-19 relevant pathway (*SARS-CoV-2 innate immunity evasion and cell-specific immune response*, identifier WP5039),and
- Retrieving the gene products for the identified pathway (proteins, genes, and metabolites) by querying the WikiPathways [23] SPARQL endpoint.
- Identifying the proteins and genes in the COVID-19 data set that are part of the gene products retrieved. Then analyzing the identified transcript and protein feature levels for the different study groups. In this step, the BridgeDB web service was used for ontology-based cross-mapping of transcript and protein identifiers from the different data sources, which use different identifiers for the same features. The overlap of the features identified in both -omics datasets is illustrated in Fig 6.

**Fig 5.**
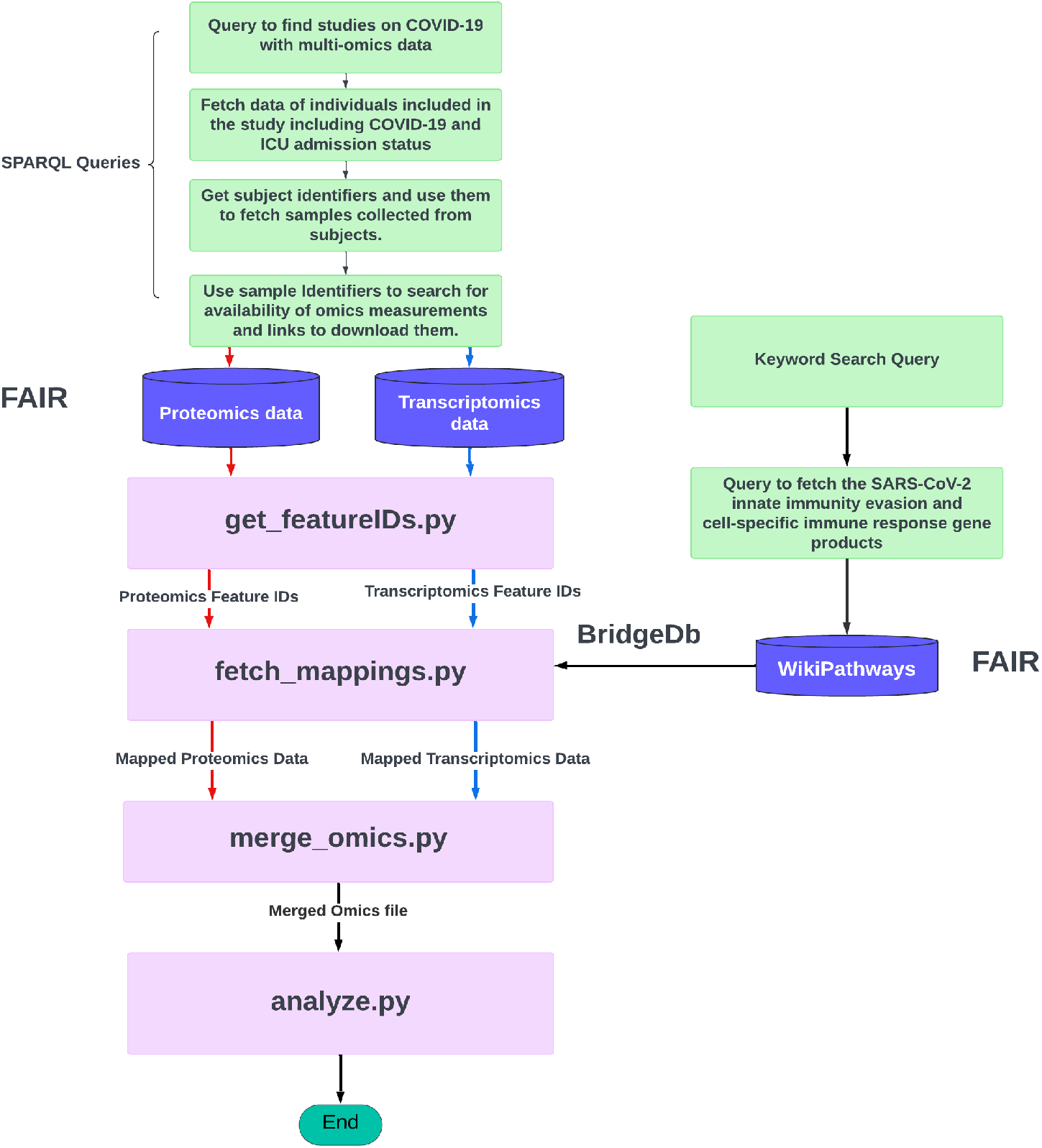
The multi-omics analysis workflow as offered in the FDCube.

**Fig 6.**
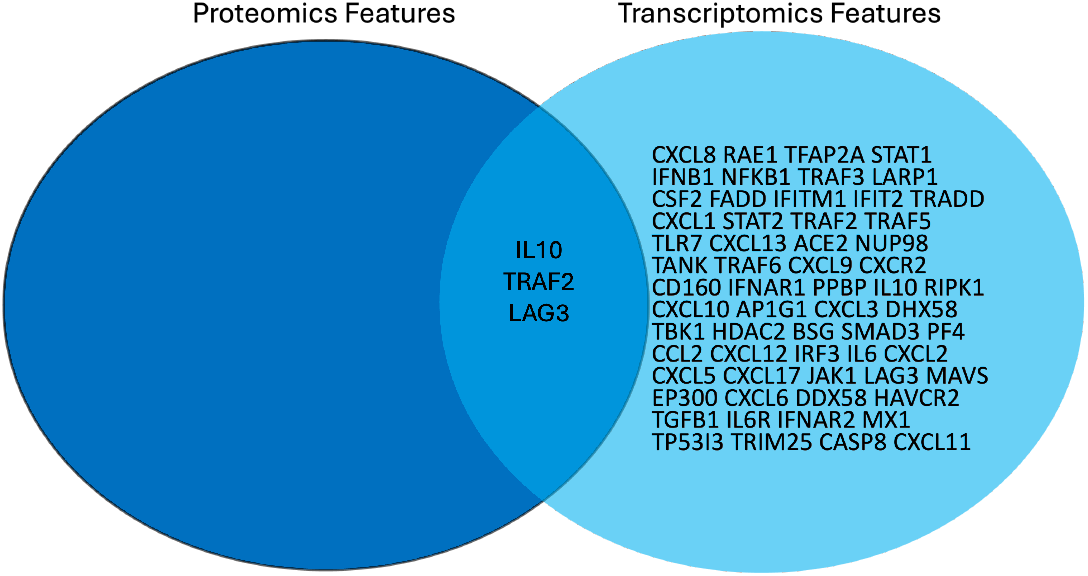
Molecular features identified in the TWOC demonstrator dataset. Overlap in proteomic and transcriptomic features as extracted from a COVID-19-relevant pathway: “*SARS-CoV-2 innate immunity evasion and cell-specific immune response*”, from WikiPathways.

One of the common features from the SARS-CoV-2 immune response pathway identified at both the transcript and protein level was Interleukine-10 (IL-10). The abundances of the transcript and protein were retrieved from the transcriptomic and proteomics datasets, together with the phenotype information of the individuals in which these abundance levels were measured. There were three groups of individuals, namely, the COVID-19 patients in the ICU, the COVID-19 patients not in the ICU and healthy individuals. The resulting box plots of IL-10 levels for these groups of individuals are presented in Fig. 7 and Fig. 8.

**Fig 7.**
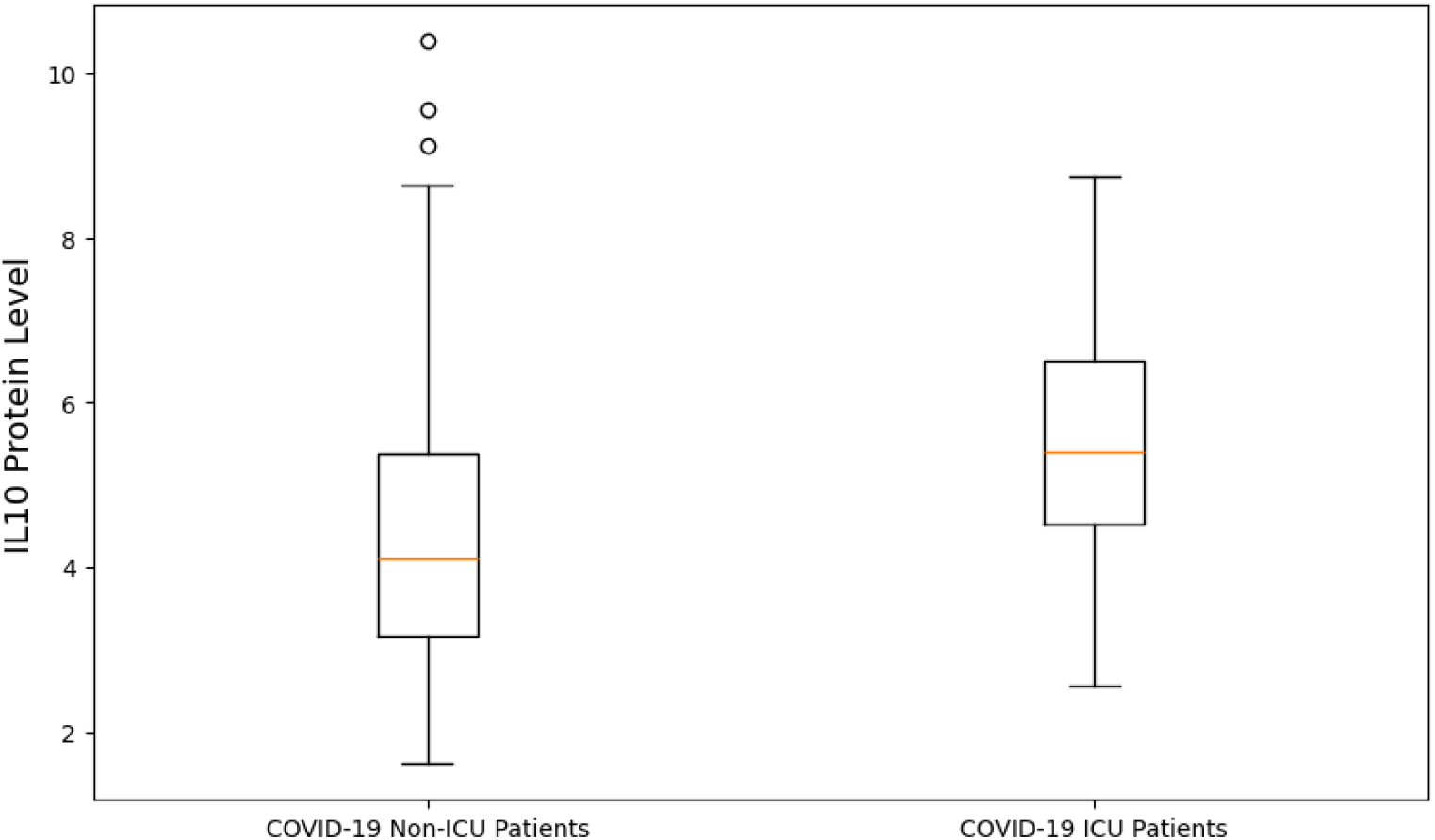
IL-10 protein level measurements for the different subject groups as identified. The y-axis represents the IL-10 protein levels, which are measured on a relative, continuous scale and indicate the concentration of the IL10 protein.

**Fig 8.**
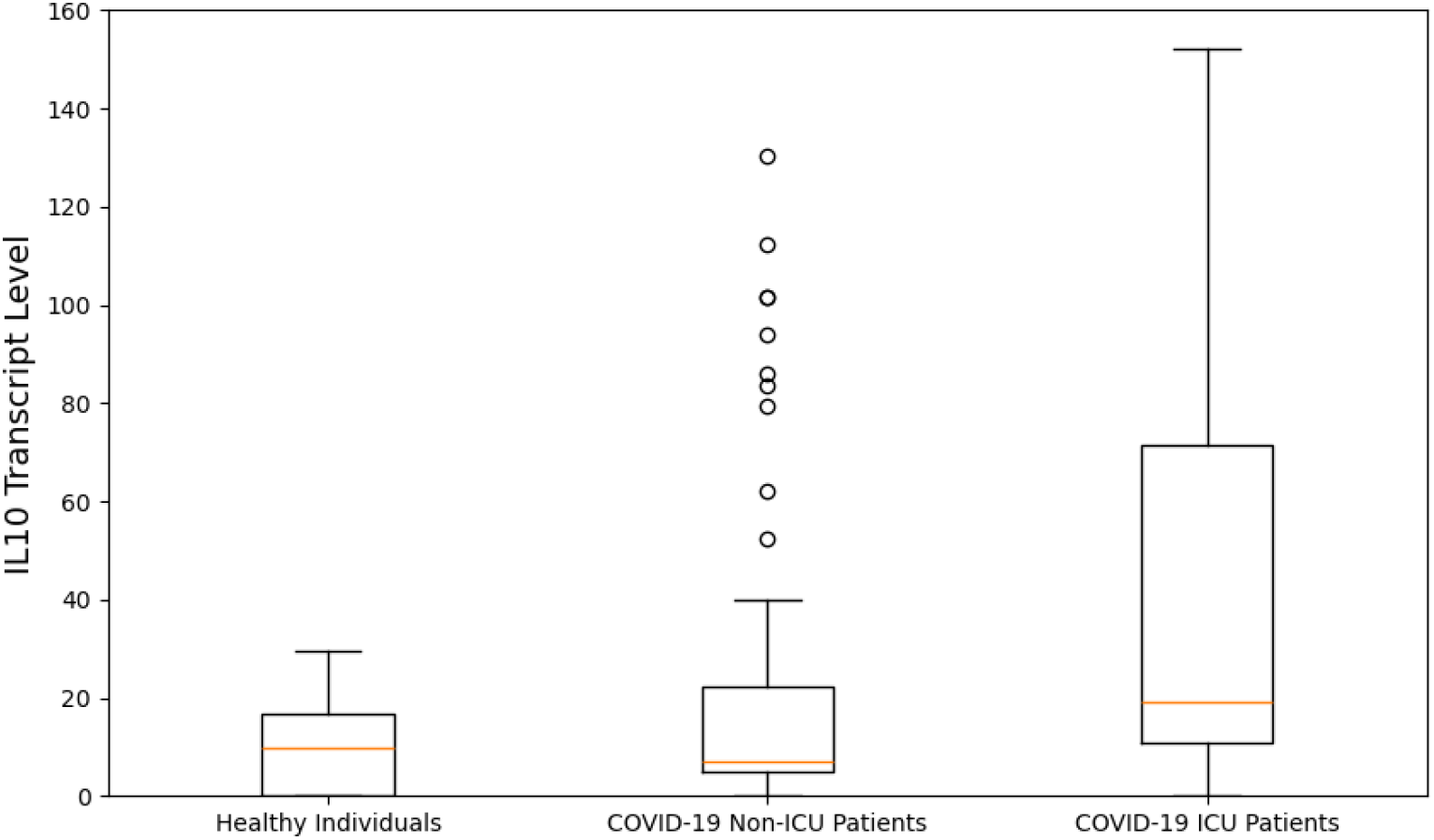
*IL10* transcript level measurements for the different subject groups as identified. The y-axis represents the *IL10* transcript levels, measured on a continuous scale, reflecting the gene expression levels of *IL10*.

The availability of FAIR data resources makes it possible to combine different data sources as shown in this multi-omics data analysis. This enables interoperability and reusability of data in a fast and efficient manner.

### 4.5 Federated analysis

This section demonstrates the federated analysis possibilities available in the FDCube, on how to deliver an algorithm to a dataset via the Vantage6 component. This example dataset is also a .*csv* file prepared from the TWOC demonstrator study. Unlike the previous example, where the dataset is publicly available on GitHub, this dataset remains in a secure environment managed by the dataset owner. The only way to access the dataset is via the help of a Vantage6 component.

A Vantage6 node is typically installed at a dataset station. For security reason, the dataset station could stay in an access-protected environment, for example, in a Digital Research Environment[34], which is a cloud based, globally available research environment.

The Vantage6 server handles authentication, keeps track of all computation requests, assigns them to nodes for computation, and stores the returning results of the analyses. The Vantage6 server could also host a private Docker registry.

Vantage6 delivers the user’s computational request to a (FAIR) data station. A computation request consists of:

- A reference to a Docker image, which contains the code (computation algorithm) that the researcher would like to run on the target dataset;
- A list describing the dataset of interest and its purpose-of-use.

Fig 9 shows the Vantage6 user interface, at which a researcher can create a task to send to the data owner(s) for federated analysis.

**Fig 9.**
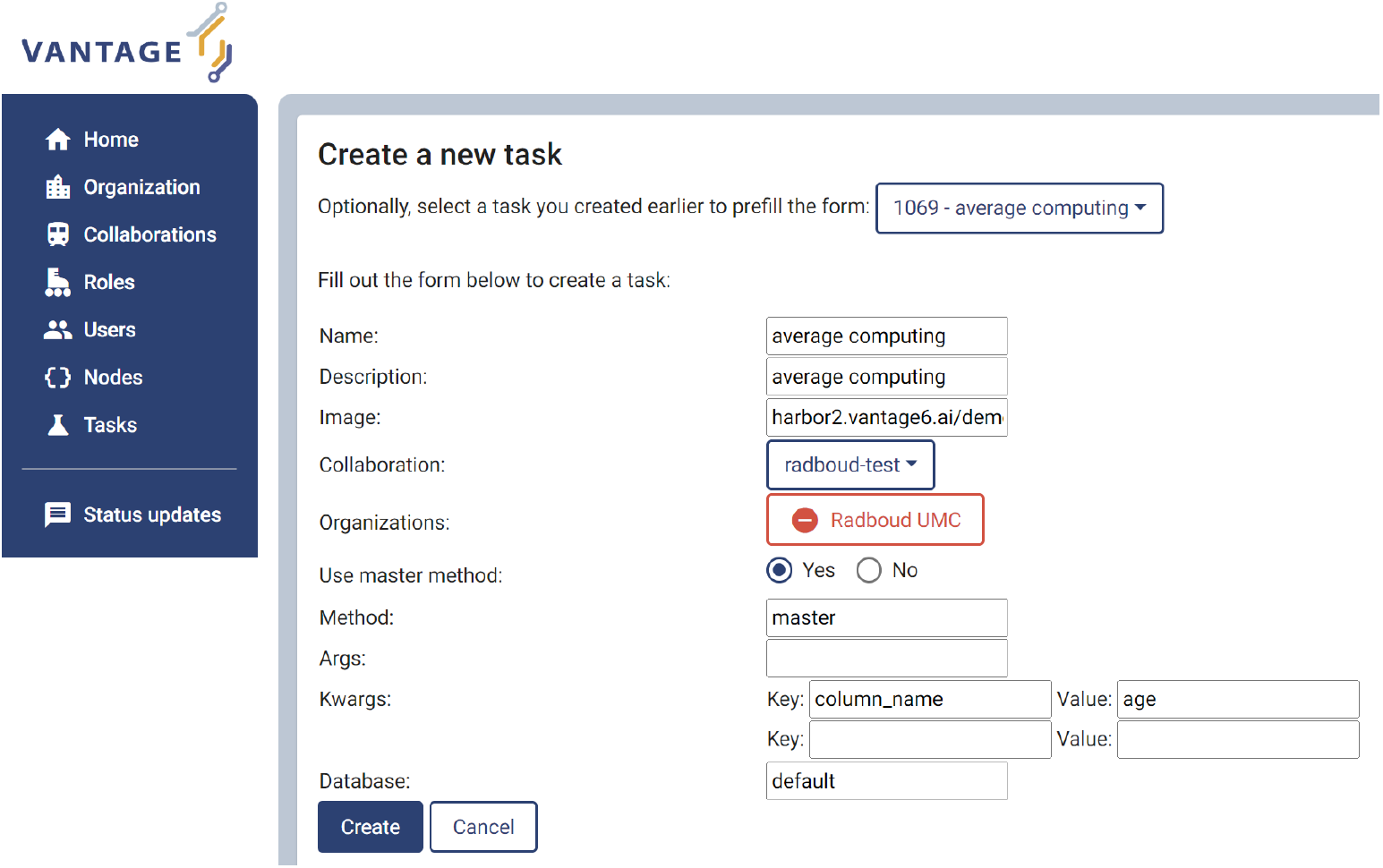
Example of creating a computation task within the Vantage6 user interface.

In this example, we used an averaging algorithm hosted on Docker Hub^1^. This algorithm expects an argument ‘*column name*’ to be defined, and will compute the average over that column. We specified in the *kwargs* fields the parameter ‘*column name*’ with value ‘*age*’. The averaging algorithm is dispatched to run on a Vantage6 node, where the dataset is stored. In this example, the dataset is a .*csv* file prepared from the FAIRified TWOC demonstrator study, which contains a column titled ‘*age*’. The ‘*Database*’ field in Fig 9 is labeled as ‘*default* ‘, which is configurable in the Vantage6 node configuration file. For simplicity, this task is created for a collaboration with only one organization (in our example: Radboudumc).

Fig 10 shows the result of running the averaging algorithm on the patients’ age in the TWOC dataset, which specifically calculates the average value in the column labelled ‘*age*’. This result can be passed back as the response to the computation request.

**Fig 10.**
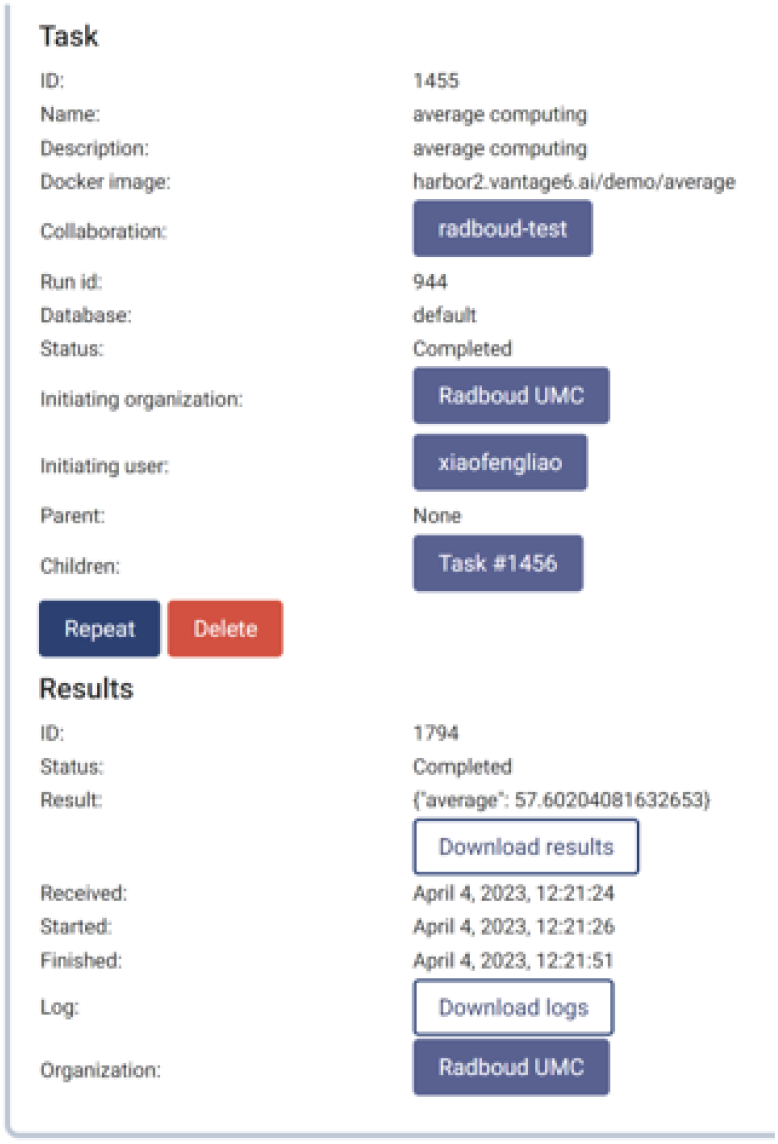
The Vantage6 portal for federated computation request, as part of the FDCube. This figure shows a snapshot of an example federated analysis task running, as displayed in the Vantage6 portal.

## 5 Conclusion

We have created the FDCube, a software and programmatic infrastructure to make (multi-)omics data FAIR, and to facilitate the management, reuse, integration and (federated) analysis of biomedical (-omics) data. The FDCube ensures data sovereignty, by utilizing Vantage6’s capability of ‘*bringing research questions to data*’ rather than ‘*sending data to research questions*’. Vantage6’s management capability covers comprehensive aspects (including organization, collaboration, users, roles, nodes and tasks), and makes FDCube a useful platform to carry out cross-organization federated analysis on decentralized datasets.

We used the FDCube in the TWOC project to demonstrate its capability and usage in creating and publishing ISA and phenotype metadata, browsing and querying the metadata on the FDP, and creating and running federated data analysis on a real dataset.

There are several ways to improve and extend the design and implementation of the current FDCube.

We are exploring the FAIR Data Station[3] for the creation of metadata, which allows a user to create a metadata template by selecting metadata fields and sheets corresponding to the user’s research, in our case, the ISA metadata schema. The metadata information captured will be ultimately transformed into a Linked Data file after a validation process.

A Beacon[35] component can be integrated into FDCube. The reason for this integration is that a FDP (by design) only exposes metadata of datasets. In contrast, Beacon allows for more insights about the content of the dataset itself, for example the presence/absence of specific genomic mutations in a set of data[35]. The combined information from both metadata (via the FDP) and real data (via Beacon), would help a researcher to get more insights into possibly available datasets, before designing a data analysis request as dictated by the researcher’s study questions.

Another potential work would be, to in the FDCube also integrate DataSHIELD with Vantage6, in order to grant users of Vantage6 access to rich analysis algorithms as available in DataSHIELD.

## Data Availability

All data produced in the present study are available upon reasonable request to the authors

https://github.com/Xomics/FAIRDataCube

## Declarations

### Funding

This work was funded by a Dutch Research Council (NWO) grant to The Netherlands X-omics Initiative (project 184.034.019), a Horizon2020 grant to the European Joint Programme on Rare Diseases (grant agreement Number 825575), a Horizon2020 grant to the EATRIS-Plus project (grant agreement Number 871096), a NWO Open Science Fund (grant agreement number 17703) and a LSH HealthHolland grant to the Trusted World of Corona (TWOC) consortium.

### Conflict of interest/Competing interests (check journal-specific guidelines for which heading to use)

No competing interest is declared.

### Ethics approval

Not applicable

### Consent to participate

Not applicable

### Consent for publication

Not applicable

### Availability of data and materials

Not applicable

### Code availability

https://github.com/Xomics/FAIRDataCube

### Authors’ contributions

P.A.C.H., A.J.G, M.A.S conceived the project. J.H. worked on phenotype data modelling. A.N., C.V worked on ISA metadata. T.E. managed connection to the TWOC project and FAIRification of the presented dataset. P.K worked on lipidomics metadata. C.D. promoted FDCube and provided scientific feedback. F.B worked on the Multi omics analysis example. Y.O implemented the FDCube as a catalog item in SURF Research Cloud. M.B supported the hosting environment. K.J.V provided insights from MOLGENIS perspective. A.N. presented the high level concept diagram. C.D. revised the Figure 2. X.L. implemented and set up the architecture of FDCube with help from all team members. X.L. wrote the manuscript with critical input and revisions from T.E., A.N., C.D., C.V., J.H., P.A.C.H, P.K., K.J.V., A.J.G. All authors reviewed the manuscript.

## Abbreviations

COVID-19: Coronavirus disease 2019
DNA: Deoxyribonucleic Acid
EMBL-EBI: EMBL’s European Bioinformatics Institute
FAIR: Findable, Accessible, Interoperable, and Reusable
FDCube: FAIR Data Cube
FDP: FAIR Data Point
ISA: Investigation, Study, Assay
PHT: Personal Health Train
RDF: Resource Description Framework
RNA: Ribonucleic Acid
SPARQL: SPARQL Protocol and RDF Query Language
TWOC: Trusted World of Corona
IL-10: Interleukine-10
OmicsDI: Omics Discovery Index

## Footnotes

1 harbor2.vantage6.ai/demo/average

## Notes

### Competing Interest Statement

The authors have declared no competing interest.

### Summary of Updates

The multi-omics analysis experiment was revised to demonstrate the FDCube capability of facilitating integrated multi-omics data analysis. Specifically to analysis IL-10 protein level measurements and IL10 transcript level measurements on data from COVID-19 patients. During this process, several FAIR resources and pathway information collected from WikiPathways were also used. New figures are generated. The axes and figure captions were more detailed. Related work was revised. New figures are added.

## References

[1] Wilkinson, M.D., Dumontier, M., Aalbersberg, I.J., Appleton, G., Axton, M., Baak, A., Blomberg, N., Boiten, J.-W., Silva Santos, L.B., Bourne, P.E., et al.: The fair guiding principles for scientific data management and stewardship. Scientific data 3(1), 1–9 (2016)

[2] Trust World of Corona. https://www.health-holland.com/project/2020/trusted-world-of-corona. Accessed: 2020-04-19

[3] Nijsse, B., Schaap, P.J., Koehorst, J.J.: Fair data station for lightweight metadata management & validation of omics studies. bioRxiv (2022) 10.1101/2022.08.03.502622 https://www.biorxiv.org/content/early/2022/08/05/2022.08.03.502622.full.pdf

[4] FiaB: FAIR-in-a-box. https://github.com/ejp-rd-vp/FiaB. Accessed: 2020-04-19

[5] DataFAIRifier. https://github.com/MaastrichtU-CDS/DataFAIRifier. Accessed: 2020-04-19

[6] Velde, K.J., Imhann, F., Charbon, B., Pang, C., Enckevort, D., Slofstra, M., Barbieri, R., Alberts, R., Hendriksen, D., Kelpin, F., et al.: Molgenis research: advanced bioinformatics data software for non-bioinformaticians. Bioinformatics 35(6), 1076–1078 (2019)

[7] Velde, K.J., Singh, G., Kaliyaperumal, R., Liao, X., Ridder, S., Rebers, S., Kerstens, H.H., Andrade, F., Reeuwijk, J., De Gruyter, F.E., et al.: Fair genomes metadata schema promoting next generation sequencing data reuse in dutch healthcare and research. Scientific data 9(1), 169 (2022)

[8] Beyan, O., Choudhury, A., Soest, J., Kohlbacher, O., Zimmermann, L., Stenzhorn, H., Karim, M.R., Dumontier, M., Decker, S., Silva Santos, L.O.B., Dekker, A.: Distributed Analytics on Sensitive Medical Data: The Personal Health Train. Data Intelligence 2(1-2), 96–107 (2020)

[9] Gaye, A., Marcon, Y., Isaeva, J., LaFlamme, P., Turner, A., Jones, E.M., Minion, J., Boyd, A.W., Newby, C.J., Nuotio, M.-L., Wilson, R., Butters, O., Murtagh, B., Demir, I., Doiron, D., Giepmans, L., Wallace, S.E., Budin-Ljøsne, I., Oliver Schmidt, C., Boffetta, P., Boniol, M., Bota, M., Carter, K.W., deKlerk, N., Dibben, C., Francis, R.W., Hiekkalinna, T., Hveem, K., Kvaløy, K., Millar, S., Perry, I.J., Peters, A., Phillips, C.M., Popham, F., Raab, G., Reischl, E., Sheehan, N., Waldenberger, M., Perola, M., Heuvel, E., Macleod, J., Knoppers, B.M., Stolk, R.P., Fortier, I., Harris, J.R., Woffenbuttel, B.H., Murtagh, M.J., Ferretti, V., Burton, P.R.: DataSHIELD: taking the analysis to the data, not the data to the analysis. International Journal of Epidemiology 43(6), 1929–1944 (2014) 10.1093/ije/dyu188 https://academic.oup.com/ije/article-pdf/43/6/1929/18482399/dyu188.pdf

[10] Moncada-Torres, A., Martin, F., Sieswerda, M., Soest, J., Geleijnse, G.: Vantage6: an open source privacy preserving federated learning infrastructure for secure insight exchange. In: AMIA Annual Symposium Proceedings, pp. 870–877 (2020)

[11] Smits, D., Beusekom, B., Martin, F., Veen, L., Geleijnse, G., Moncada-Torres, A.: An improved infrastructure for privacy-preserving analysis of patient data. In: Proceedings of the International Conference of Informatics, Management, and Technology in Healthcare (ICIMTH), vol. 295, pp. 144–147 (2022)

[12] R Core Team: R: A Language and Environment for Statistical Computing. R Foundation for Statistical Computing, Vienna, Austria (2021). R Foundation for Statistical Computing. https://www.R-project.org/

[13] Research & Innovation., E.C.D.-G.: H2020 programme guidelines on fair data management in horizon 2020 (2016)

[14] Silva Santos, L.O.B., Burger, K., Kaliyaperumal, R., Wilkinson, M.D.: FAIR Data Point: A FAIR-Oriented Approach for Metadata Publication. Data Intelligence, 1–21 (2022) 10.1162/dinta00160

[15] Musen, M.A., Bean, C.A., Cheung, K.-H., Dumontier, M., Durante, K.A., Gevaert, O., Gonzalez-Beltran, A., Khatri, P., Kleinstein, S.H., O’Connor, M.J., Pouliot, Y., Rocca-Serra, P., Sansone, S.-A., Wiser, J.A.,, CEDAR team: The center for expanded data annotation and retrieval. Journal of the American Medical Informatics Association 22(6), 1148–1152 (2015) 10.1093/jamia/ocv048 https://academic.oup.com/jamia/article-pdf/22/6/1148/34146117/ocv048.pdf

[16] Wolstencroft, K., Krebs, O., Snoep, J.L., Stanford, N.J., Bacall, F., Golebiewski, M., Kuzyakiv, R., Nguyen, Q., Owen, S., Soiland-Reyes, S., Straszewski, J., van Niekerk, D.D., Williams, A.R., Malmströom, L., Rinn, B., Muöller, W., Goble, C.: FAIRDOMHub: a repository and collaboration environment for sharing systems biology research. Nucleic Acids Research 45(D1), 404–407 (2016) 10.1093/nar/gkw1032 https://academic.oup.com/nar/article-pdf/45/D1/D404/8846631/gkw1032.pdf

[17] Perez-Riverol, Y., Bai, M., Veiga Leprevost, F., Squizzato, S., Park, Y.M., Haug, K., Carroll, A.J., Spalding, D., Paschall, J., Wang, M., et al.: Discovering and linking public omics data sets using the omics discovery index. Nature biotechnology 35(5), 406–409 (2017)

[18] Sansone, S.A., Rocca Serra, P., Field, D., Maguire, E., Taylor, C., Hofmann, O., Fang, H., Neumann, S., Tong, W., Amaral Zettler, L., Begley, K., Booth, T., Bougueleret, L., Burns, G., Chapman, B., Clark, T., Coleman, L.A., Copeland, J., Das, S., de Daruvar, A., de Matos, P., Dix, I., Edmunds, S., Evelo, C.T.A., Forster, M.J., Gaudet, P., Gilbert, J., Goble, C., Griffin, J.L., Jacob, D., Kleinjans, J., Harland, L., Haug, K., Hermjakob, H., Ho Sui, S.J., Laederach, A., Liang, S., Marshall, S., McGrath, A., Merrill, E., Reilly, D., Roux, M., Shamu, C.E., Shang, C.A., Steinbeck, C., Trefethen, A., Jones, B., Wolstencroft, K., Xenarios, I., Hide, W.: Toward interoperable bioscience data. Nature Genetics 44(2), 121–126 (2012) 10.1038/ng.1054

[19] Johnson, D., Batista, D., Cochrane, K., Davey, R.P., Etuk, A., Gonzalez-Beltran, A., Haug, K., Izzo, M., Larralde, M., Lawson, T.N., Minotto, A., Moreno, P., Nainala, V.C., O’Donovan, C., Pireddu, L., Roger, P., Shaw, F., Steinbeck, C., Weber, R.J.M., Sansone, S.-A., Rocca-Serra, P.: ISA API: An open platform for interoperable life science experimental metadata. GigaScience 10(9) (2021) 10.1093/gigascience/giab060 https://academic.oup.com/gigascience/article-pdf/10/9/giab060/40394493/giab060 reviewer 3 report revision 1.pdf. giab060

[20] Ladewig, M.S., Jacobsen, J.O.B., Wagner, A.H., Danis, D., El Kassaby, B., Gargano, M., Groza, T., Baudis, M., Steinhaus, R., Seelow, D., Bechrakis, N.E., Mungall, C.J., Schofield, P.N., Elemento, O., Smith, L., McMurry, J.A., Munoz-Torres, M., Haendel, M.A., Robinson, P.N.: Ga4gh phenopackets: A practical introduction. Advanced Genetics n/a(n/a), 2200016 10.1002/ggn2.202200016 https://onlinelibrary.wiley.com/doi/pdf/10.1002/ggn2.202200016

[21] MetaboLights. https://www.ebi.ac.uk/metabolights/. Accessed: 2020-04-19

[22] Su, Y., Chen, D., Yuan, D., Lausted, C., Choi, J., Dai, C.L., Voillet, V., Duvvuri, V.R., Scherler, K., Troisch, P., Baloni, P., Qin, G., Smith, B., Kornilov, S.A., Rostomily, C., Xu, A., Li, J., Dong, S., Rothchild, A., Zhou, J., Murray, K., Edmark, R., Hong, S., Heath, J.E., Earls, J., Zhang, R., Xie, J., Li, S., Roper, R., Jones, L., Zhou, Y., Rowen, L., Liu, R., Mackay, S., O’Mahony, D.S., Dale, C.R., Wallick, J.A., Algren, H.A., Zager, M.A., Wei, W., Price, N.D., Huang, S., Subramanian, N., Wang, K., Magis, A.T., Hadlock, J.J., Hood, L., Aderem, A., Bluestone, J.A., Lanier, L.L., Greenberg, P.D., Gottardo, R., Davis, M.M., Goldman, J.D., Heath, J.R.: Multi-omics resolves a sharp disease-state shift between mild and moderate covid-19. Cell 183(6), 1479–149520 (2020) 10.1016/j.cell.2020.10.037

[23] Agrawal, A., Balcı, H., Hanspers, K., Coort, S.L., Martens, M., Slenter, D.N., Ehrhart, F., Digles, D., Waagmeester, A., Wassink, I., Abbassi-Daloii, T., Lopes, E.N., Iyer, A., Acosta, J., Willighagen, L.G., Nishida, K., Riutta, A., Basaric, H., Evelo, C., Willighagen, E.L., Kutmon, M., Pico, A.: WikiPathways 2024: next generation pathway database. Nucleic Acids Research 52(D1), 679–689 (2023) 10.1093/nar/gkad960 https://academic.oup.com/nar/article-pdf/52/D1/D679/55040703/gkad960.pdf

[24] TWOC demonstrator. https://github.com/Xomics/TWOCdemonstrator/tree/main/data/Su 2020 original/phenotypes in modules. Accessed: 2020-04-19

[25] Rocca-Serra, P., Maguire, E., Taylor, C., Field, D., Wittenberger, T., Santarsiero, A., Gonzalez-Beltran, A., Sansone, S.-A.: 7 - investigation-study-assay, a toolkit for standardizing data capture and sharing. In: Harland, L., Forster, M. (eds.) Open Source Software in Life Science Research. Woodhead Publishing Series in Biomedicine, pp. 173–188. Woodhead Publishing, ??? (2012). 10.1533/9781908818249.173. https://www.sciencedirect.com/science/article/pii/B9781907568978500072

[26] RDF 1.1 Turtle. http://www.w3.org/TR/2014/REC-turtle-20140225/

[27] TWOC Demonstrator Tools. https://github.com/Xomics/TWOCdemonstrator/tree/main/tools. Accessed: 2020-04-19

[28] Heyvaert, P., De Meester, B., Dimou, A., Verborgh, R.: Declarative rules for linked data generation at your fingertips! In: Gangemi, A., Gentile, A.L., Nuzzolese, A.G., Rudolph, S., Maleshkova, M., Paulheim, H., Pan, J.Z., Alam, M. (eds.) The Semantic Web: ESWC 2018 Satellite Events, pp. 213–217. Springer, Cham (2018)

[29] Phenopackets RDF Sschema. https://github.com/LUMC-BioSemantics/phenopackets-rdf-schema. Accessed: 2020-04-19

[30] ISA tools API. https://isa-tools.org/isa-api/content/index.html. Accessed: 2020-04-19

[31] ISA tools environment. https://github.com/Xomics/Isatoolsenvironment. Accessed: 2020-04-19

[32] The FAIR Data Point in CMBI. https://fdp.cmbi.umcn.nl. Accessed: 2020-04-19

[33] TWOC Demonstrator Interleukine-6 (IL-6) Analysis. https://github.com/Xomics/TWOCdemonstrator/blob/main/tools/pythonreadomics/IL6.ipynb. Accessed: 2024-05-07

[34] Digital Research Environment. https://www.radboudumc.nl/en/research/radboud-technology-centers/data-stewardship/digital-research-environment. Accessed: 2020-04-19

[35] Rambla, J., Baudis, M., Ariosa, R., Beck, T., Fromont, L.A., Navarro, A., Paloots, R., Rueda, M., Saunders, G., Singh, B., Spalding, J.D., Töornroos, J., Vasallo, C., Veal, C.D., Brookes, A.J.: Beacon v2 and beacon networks: A “lingua franca” for federated data discovery in biomedical genomics, and beyond. Human Mutation 43(6), 791–799 (2022) 10.1002/humu.24369 https://onlinelibrary.wiley.com/doi/pdf/10.1002/humu.24369

